# Comprehensive Evaluation of Mosquito Repellent Products: Efficacy, Safety, and Public Health Implications

**DOI:** 10.64898/2026.05.07.26352623

**Authors:** Md Ushama Shafoyat, Tamanna Mostafa, Kazi Sahal, Biprodab Mogumdar, kaiissar mannoor, Shahriar Kabir Sium, SM Al-Amin

## Abstract

Mosquito-borne diseases such as dengue, chikungunya, and malaria remain major public health challenges in South Asia, particularly in Bangladesh, where mosquito repellents are widely used as primary preventive tools. This study presents a comparative evaluation of commonly used repellent products, including mosquito coils, liquid vaporizers, aerosols, DEET-based creams, and natural formulations, focusing on their efficacy, duration of protection, and potential health risks. Efficacy was assessed using controlled laboratory methods, including chamber-based exposure and arm-in-cage tests against Aedes aegypti. Safety was evaluated in vivo using a rat model through clinical observations, hematological and biochemical profiling, and histopathological examination. The results indicated an overall mean effectiveness of 85%, with DEET-based creams demonstrating the highest efficacy (95%) and natural repellents the lowest (70%). Mosquito coils provided the longest protection duration (∼10 hours) but posed the most severe health hazards due to combustion-related emissions. Toxicological findings revealed that prolonged coil exposure induced significant physiological damage, including reduced body weight gain, elevated leukocyte counts, decreased hemoglobin, altered liver and kidney biochemical markers, and severe histopathological damage in lung, liver, and skin tissues. Vaporizers and aerosols resulted in comparatively milder physiological changes. In contrast, DEET-based products and natural formulations demonstrated negligible systemic toxicity, preserving normal organ structure and function. Overall, the study highlights a clear efficacy–safety trade-off. Based on these findings, vaporizers or aerosols are preferred for ventilated indoor use, DEET-based repellents are recommended for prolonged outdoor activities, and natural formulations serve as ideal alternatives for vulnerable populations (children, pregnant women, the elderly, and those with respiratory sensitivities). These results underscore the need for informed consumer selection, integrated vector management strategies, stronger regulatory oversight, and the development of sustainable, low-toxicity mosquito-repellent technologies.

## 1. Introduction

Mosquito-borne diseases continue to impose a substantial public health burden across South Asia, with recurrent outbreaks of dengue, chikungunya, and malaria disproportionately affecting densely populated urban and peri-urban settings (Peng et al., 2022a). Rapid urbanization, inadequate waste management, and favorable climatic conditions have intensified the proliferation of *Aedes* mosquitoes, particularly *Aedes aegypti*, which is the primary vector of dengue. In countries such as Bangladesh, seasonal epidemics have become increasingly frequent and severe, placing significant strain on healthcare systems and highlighting the urgent need for effective preventive strategies (Katz et al., 2008).

In the absence of universally accessible vaccines and specific antiviral therapies for dengue, personal protection and vector control remain the cornerstone of disease prevention. Among these, mosquito repellent products are extensively used at the household level in a variety of formulations. The Bangladeshi market offers a wide range of commercially available repellents, including mosquito coils such as (Nguyen et al., 2023) RAK Mosfly and Eagle Max Hit, liquid vaporizers such as Good Knight and Mortein, aerosol sprays marketed under brands like ACI Aerosol, and topical formulations including DEET-based creams, Dabur Odomos cream (Chen-Hussey et al., 2014) and plant-derived repellents such as Leeings mosquito repellent and Mamaearth Natural Mosquito Repellent Spray (Maia & Moore, 2011; Webb & Hess, 2016). These products differ significantly in their active ingredients, modes of action, duration of protection, and routes of human exposure (Katz et al., 2008).

Market analyses indicate that mosquito coils dominate consumer usage, accounting for approximately 88% of repellent consumption, primarily due to their low cost, widespread availability, and ease of use. Despite their popularity, mosquito coils and other combustion-based repellents have raised concerns regarding indoor air pollution and associated health risks (Nguyen et al., 2023), including respiratory irritation and long-term exposure to particulate matter and volatile organic compounds (Peterson et al., 2006). Similarly, while liquid vaporizers and aerosol sprays offer greater convenience and reduced smoke emissions, their reliance on synthetic pyrethroids raises potential toxicological concerns. Topical repellents, particularly those containing N,N-diethyl-meta-toluamide (DEET) (Chen-Hussey et al., 2014), are widely regarded as effective; however, issues related to dermal absorption, misuse, and public perception persist (Nguyen et al., 2023). Natural or plant-based repellents are increasingly favored for their perceived safety profile, yet their cost and limited availability make them difficult to find in mosquito-affected areas, and market syndicates narrow their supply (Maia & Moore, 2011; Webb & Hess, 2016).

Given the heterogeneity of products and the lack of standardized performance and safety evaluations in local markets, there is a critical need for a systematic, comparative assessment of mosquito repellent technologies. Existing studies often focus on individual product categories or controlled laboratory conditions, with limited integration of real-world usage patterns and exposure risks (Maia & Moore, 2011). Furthermore, inconsistencies in product quality and regulatory oversight in developing markets exacerbate uncertainties regarding both efficacy and safety (Diaz, 2016). Therefore, this study aims to provide a comprehensive evaluation of commonly used mosquito repellent products in Bangladesh by integrating laboratory-based efficacy testing, toxicological assessment, and field-level performance analysis (Diaz, 2016). The findings are expected to inform evidence-based consumer choices, guide regulatory policies, and support the development of safer and more effective vector control interventions in dengue-endemic regions.

## 2. Overview of Common Mosquito Repellent Products in South Asia

Mosquito repellent products available in Bangladesh encompass a diverse range of formulations designed for both environmental and personal protection. These products differ in their active ingredients, mode of delivery, duration of protection, and safety profiles. The most commonly used categories include mosquito coils, liquid vaporizers, aerosols, and natural repellents (Shyam et al., 2026), and chemical (DEET-based) topical creams (Chen-Hussey et al., 2014; Sudakin & Trevathan, 2003; Webb & Hess, 2016).

Mosquito coils remain the most widely used repellent product in Bangladesh due to their affordability, ease of use, and accessibility across both urban and rural settings (Liu et al., 2024). Commonly available brands include RAK Mosfly, Eagle Max Hit, FINIS Black Booster, Bashundhara Xtreme, Rak Super Mosfly, Baoma, Pata Bahar, Neempata, and Dragon, as well as locally marketed products such as Zero Bite and Sweet Dream. These coils typically contain synthetic pyrethroid compounds, primarily D-trans allethrin and dimefluthrin, which act as neurotoxic agents to mosquitoes, leading to knockdown and repellency (Liu et al., 2024). When ignited, coils release carbon dioxide and carbon monoxide, along with insecticidal smoke that disperses into the surrounding environment, reducing the oxygen percentage in closed rooms. In terms of performance, mosquito coils can provide approximately 10–12 hours of protection per unit, making them suitable for overnight use (Liu et al., 2024). They are effective in both indoor and semi-open outdoor environments. However, their efficacy is influenced by factors such as ventilation, room size, and air circulation (Diaz, 2016).

Liquid vaporizers and aerosols represent a modern alternative to coils, offering a smoke-free and more controlled method of mosquito repellency. Popular brands in Bangladesh include Good Knight, ACI Aerosol, and Mortein. These devices operate by electrically heating a liquid formulation containing pyrethroid-based active ingredients, which then evaporate into the air. Vaporizers and aerosols are designed for use in enclosed indoor environments and typically provide continuous protection throughout the night. A single refill unit can last approximately 45–60 nights, depending on usage duration, making them a convenient long-term option for households. The absence of smoke emission reduces indoor particulate pollution compared to coils; however, their effectiveness is limited to enclosed spaces and depends on proper device functioning and room conditions (Liu et al., 2024).

Natural or plant-based repellents have gained increasing attention due to their perceived safety and environmental friendliness (Maia & Moore, 2011; Nguyen et al., 2023). Examples available in the Bangladeshi market include Leeings mosquito repellent liquid, Mamaearth Natural Mosquito Repellent Spray, citronella-based spray, and Leeings natural oil-based formulations used with humidifiers. These products are typically free from synthetic chemicals and rely on essential oils, including olive oil, thyme oil, clove oil, citronella oil, lemongrass oil, eucalyptus oil, chrysanthemum oil, and peppermint, for their repellent action (Chen-Hussey et al., 2014). The mechanism involves masking human odor cues and interfering with mosquito host-seeking behavior. Natural repellents (Leeings mosquito repellent and Mamaearth Natural Mosquito Repellent Spray) are generally considered safe for sensitive populations, including children and individuals with respiratory concerns. However, their effective protection time depends on 4-6 hours in enclosed spaces and on proper device functioning and room conditions

Topical mosquito repellent creams containing N, N-diethyl-meta-toluamide (DEET) are widely recognized for their high efficacy in personal protection. Products such as Dabur Odomos cream are commonly used in Bangladesh (Sudakin & Trevathan, 2003; Webb & Hess, 2016). DEET-based formulations provide good protection, typically lasting 4 to 8 hours, depending on concentration and environmental conditions. Concentrations of 20% or higher are generally considered highly effective against *Aedes* mosquitoes. These creams function by altering mosquito olfactory receptor responses, thereby preventing host detection and reducing biting incidence. While DEET is considered safe when used as directed, concerns remain regarding excessive application, dermal absorption, and potential irritation, particularly in sensitive individuals (Chen-Hussey et al., 2014).

Overall, the South Asia mosquito repellent market is characterized by a wide spectrum of products with varying levels of efficacy, safety, and user convenience (Table 1). Mosquito coils dominate usage due to economic factors, whereas vaporizers/ aerosol and DEET-based creams offer improved performance and safety profiles. Natural (Leeings mosquito repellent and Mamaearth Natural Mosquito Repellent Spray) repellents provide a low-toxicity alternative but are limited by enclosed spaces, proper device functioning, and room conditions. This diversity underscores the need for systematic evaluation to guide optimal product selection and public health recommendations (Diaz, 2016) (Peterson et al., 2006).

**Table 1:**
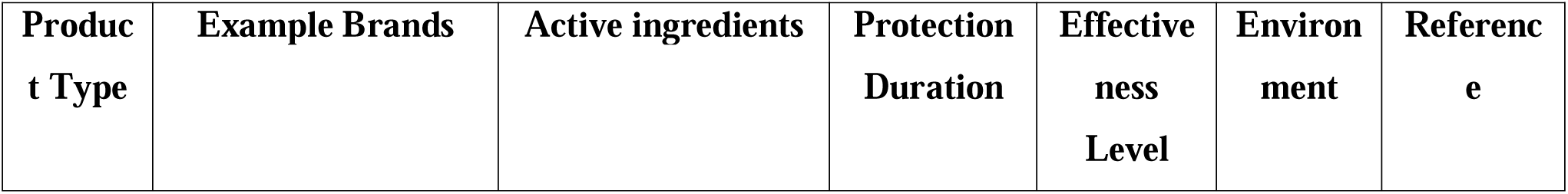
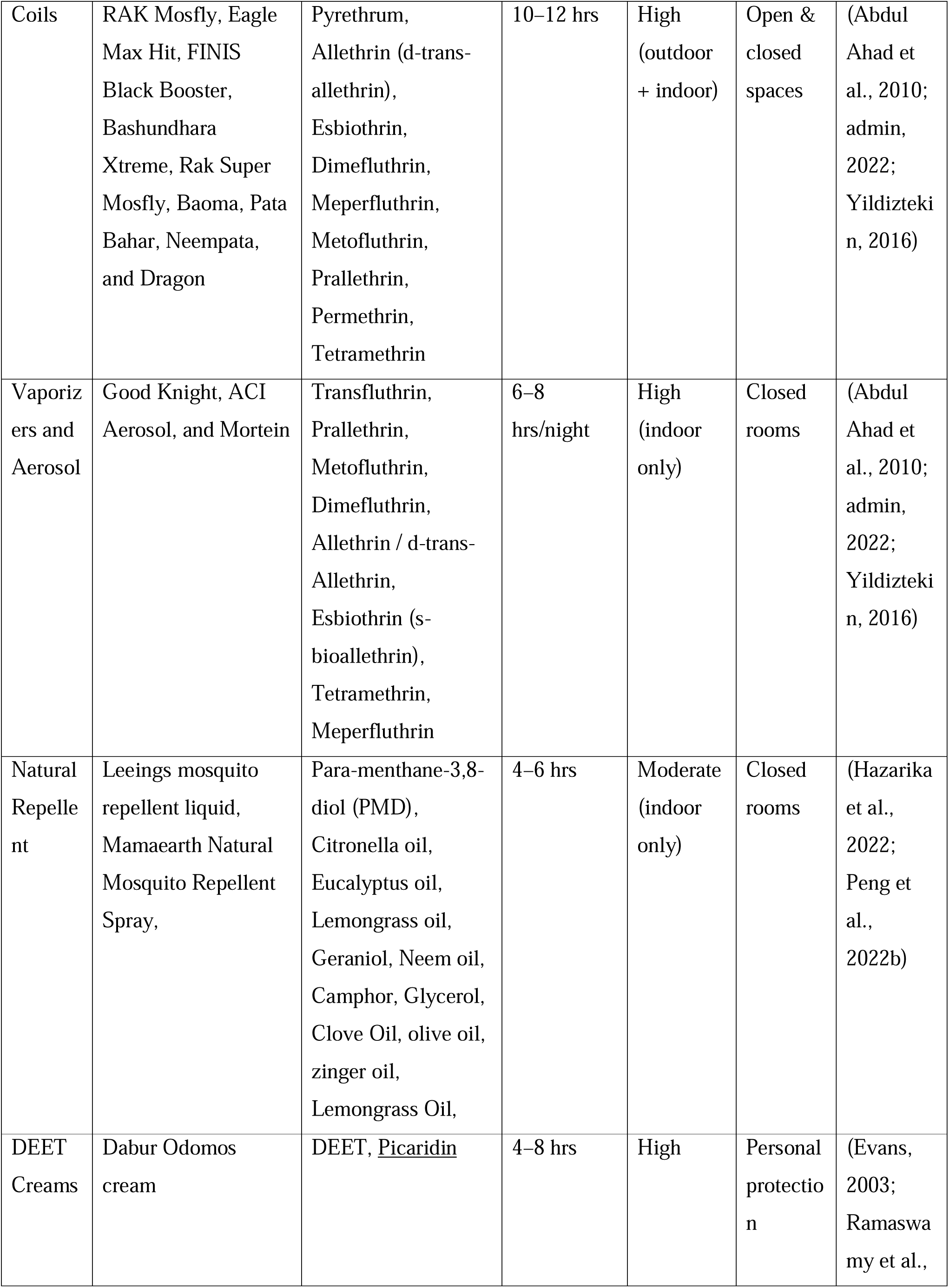
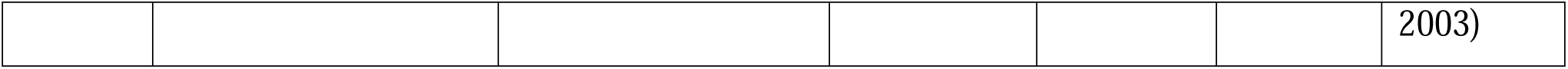
Comparative Analysis of Effectiveness based on the literature.

## 3. Materials and Methode

### 3.1 Mosquitoes Used in Repellent Tests

Aedes aegypti mosquitoes were used for repellent testing. Mosquito larvae were obtained from the IRES (Insect Rearing and Experimental Station) Department of Zoology, Jahangir Nagar University, Savar, Bangladesh. The mosquitoes were reared and tested in a climate-controlled chamber maintained at 27 ± 2°C and 70–80% relative humidity at a dedicated facility of the Military Institute of Science and Technology, conditions optimized for vector activity and survival. The experimental population specifically consisted of active, host-seeking female Aedes aegypti mosquitoes aged 5–7 days. Prior to starting the efficacy assays, the adult mosquitoes were starved for 12 hours to enhance host-seeking responsiveness and ensure consistent biting pressure. Adult mosquitoes were otherwise fed and maintained on a 10% sucrose solution, as described previously. (Gerberg et al., 1994)

### 3.2. Repellent Testing and Formulations

Four distinct categories of commercially available 15 locally available Bangladeshi market mosquito repellents were evaluated in this study to determine their comparative efficacy: coils, vaporizers/aerosols, natural liquid repellents, and topical creams. The representative test products included coils (RAK Mosfly Coil, Eagle Max Hit Coil, FINIS Black Booster, Bashundhara Xtreme, Rak Super, Baoma, Pata Bahar, Neempata, and Dragon) containing active ingredients such as Pyrethrum, Allethrin derivatives, Dimefluthrin, Meperfluthrin, Metofluthrin, Prallethrin, Permethrin, or Tetramethrin; vaporizers and aerosols (Good Knight, Mortein, and ACI Aerosol) formulated with Transfluthrin, Prallethrin, Metofluthrin, Dimefluthrin, Allethrin variations, Esbiothrin, Tetramethrin, or Meperfluthrin; natural repellents (Leeings mosquito repellent liquid and Mamaearth Natural Mosquito Repellent Spray) composed of plant-based essential oils including Citronella, Lemongrass, Neem, Eucalyptus, Clove, Ginger, onion, chrysanthemum, cardamom, and olive oil; and topical creams (Dabur Odomos cream) utilizing DEET or Picaridin as primary active protection agents. (Logan et al., 2010)

### 3.3. Test Cage Design and Construction

A custom test cage measuring 40 × 50 × 40 cm was constructed for the arm-in-cage exposure assays. To facilitate thorough decontamination between experimental trials, the enclosure was built utilizing a sturdy metal frame. All sides of the cage were fitted with a visible white mesh netting, which allowed researchers to monitor, count, and record mosquito landing and biting rates in real time. A flexible fabric sleeve was securely attached to the front panel of the cage, providing a sealed access point for the controlled introduction of the volunteer’s treated or untreated forearm into the environment containing 50 unfed female *Aedes aegypti* mosquitoes.

### 3.4. Arm-in-Cage Test Procedure

The mosquito repellent efficacy testing followed standardized methodologies adapted from the World Health Organization (WHO) guidelines for efficacy testing of mosquito repellents on human skin. For each evaluation, a controlled population of 50 unfed female *Aedes aegypti* mosquitoes that had never received a blood meal was maintained in each custom test cage. To optimize host-seeking responsiveness and ensure uniform biting pressure, these mosquitoes were starved of their regular 10% sucrose solution for 12 hours prior to the initiation of the trials.

Healthy adult volunteers (n = 6) were recruited for the study following ethical approval and informed consent. The experiment was approved by the Institutional Review Board of the Biomedical Research and Innovation Center, Military Institute of Science and Technology, Mirpur-1216, Dhaka, Bangladesh (approval code: 2025H009, Date of Approval: 25/ 09/2025, NCT07686159). The recruitment of study participants was conducted from 25/09/2025 to 29/01/2026, during which eligible volunteers were enrolled after screening against predefined inclusion and exclusion criteria, and informed consent procedures were conducted in accordance with the study protocol.

Prior to testing, each participant’s forearm was thoroughly washed with unscented soap and then allowed to air-dry for 5 minutes to eliminate any confounding odors that could influence mosquito attraction. To standardize testing across candidates, the respective formulations were applied uniformly to the volunteers’ forearms at a dose of 1 mL per 600 cm² of exposed skin before exposure to cages containing active, host-seeking mosquitoes. The treated forearm was subsequently introduced into a mosquito cage containing 50 unfed female *Aedes aegypti* mosquitoes. Volunteers were not permitted to remove or wet the patch during this time. After 8 hours, the patch was removed by medical personnel, and initial results were determined.

During each exposure session, the following parameters were recorded:

- Number of mosquito landings
- Number of confirmed bites

Efficacy Calculation:

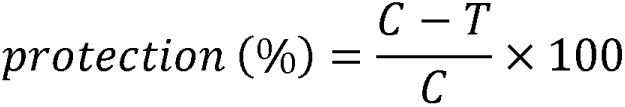

Where:

- C = bites on control arm
- T = bites on treated arm

The complete protection time (CPT) was defined as the time the first mosquito landed on or bit a treated arm. To determine the CPT of mosquito repellents, the treated right arm of each volunteer was inserted into the test cage for 3 min. If there were no bites, that arm was reinserted at 10 min intervals until the first bite occurred.

### 3.5. coils, aerosols and liquid vaporizers, and natural repellents Testing Procedure

The efficacy of mosquito coils, aerosols, and liquid vaporizers, and natural repellents was evaluated under simulated indoor conditions in a standardized, enclosed-chamber model measuring 1.8x 1.8 x 1.8 m (with a total volume of approximately 6 m^3^). The experimental setup was designed to replicate a typical residential room environment to assess both immediate and residual effects of the products. Each test product was placed and operated within an enclosed chamber of known volume under controlled environmental conditions. Following system stabilization, 100 adult female *Aedes aegypti* mosquitoes were released into the chamber and allowed to acclimatize briefly before exposure (Barnard & Xue, n.d.; Diaz, 2016). Efficacy assessment was conducted through two primary outcome measures:

- **Knockdown rate:** The number of immobilized or incapacitated mosquitoes was recorded at 10-minute intervals during the exposure period. Knockdown was defined as the inability of mosquitoes to maintain normal flight or movement.
- **Mortality rate:** After completion of the exposure period, mosquitoes were transferred to holding containers and maintained under standard conditions. Final mortality was recorded after 24 hours, with mosquitoes showing no movement considered dead.

All experiments were performed in replicates to ensure data reliability. This protocol enabled the evaluation of both the rapid action (knockdown effect). It delayed the onset of lethality (mortality) in coil, aerosol, and vaporizer formulations, as well as in natural repellent formulations, providing a comprehensive assessment of their insecticidal performance.

### 3.6. Safety and Toxicological Evaluation

An in vivo toxicological assessment was conducted using a rat model to evaluate the potential health effects of commonly used mosquito repellent products (Peterson et al., 2006). The study focused on representative formulations, including mosquito coil emissions, liquid vaporizer and aerosol exposure, DEET-based topical application, and natural repellent formulations (Chen-Hussey et al., 2014; Magesh et al., 2018; Webb & Hess, 2016). The experimental protocol was designed to simulate realistic exposure scenarios relevant to human use patterns.

#### 3.6.1. Experimental Animals

Healthy adult Wistar rats (*Rattus norvegicus*) weighing 180–220 g were used in the study. Animals were housed under controlled laboratory conditions (temperature: 22 ± 2°C; relative humidity: 50– 60%; 12-hour light/dark cycle) with free access to standard feed and water. All experimental procedures were conducted in accordance with institutional ethical guidelines and approved by the Ethical Committee (Approval Number: 2025H005; Date: 25 Jun 2025).

#### 3.6.2. Grouping and Exposure Protocol

Animals were randomly divided into five groups (A total of 24 Wistar rats, n = 3 per group):

- **Group I** (Control): No exposure
- **Group II** (Coil Exposure): Exposure to mosquito coil (RAK Mosfly Coil, Bashundhara Xtreme) smoke (8 hours/day)
- **Group III (**Vaporizer and aerosol Exposure): Exposure to vaporizer and aerosol emissions (Good night, ACI Aerosol) (8 hours/day)
- **Group IV** (DEET Application): Topical application of DEET-based cream (Dabur Odomos cream), standard dermal dose of 1 mL per 600 cm² of exposed skin (8 hours/day)
- **Group V** (Natural Repellent): Exposure using a humidifier and spray (Leeings mosquito repellent liquid, Mamaearth Natural Mosquito Repellent Spray) (8 hours/day)

The exposure duration was maintained for 28 consecutive days to assess sub-chronic toxicity.

#### 3.6.3. Ethical Considerations, Anesthesia, and Humane Endpoints

All animal experiments were conducted in accordance with institutional ethical guidelines and internationally accepted principles for the care and use of laboratory animals.

(a) Methods of sacrifice

At the end of the experimental period, animals were humanely sacrificed under deep anesthesia to minimize pain and distress. Euthanasia was performed by overdose administration of ketamine– xylazine followed by cervical dislocation to ensure rapid and irreversible death prior to tissue collection. This method was selected in accordance with standard laboratory animal welfare recommendations. (Animals, 1992)

(b) Methods of anesthesia and/or analgesia

For procedures involving handling and sample collection, animals were anesthetized using an intraperitoneal injection of Ketamine (80–100 mg/kg body weight) combined with Xylazine (5–10 mg/kg body weight) to ensure adequate anesthesia, analgesia, and muscle relaxation during experimental procedures. The depth of anesthesia was monitored by assessing pedal withdrawal and corneal reflexes prior to invasive manipulations.

(c) Efforts to alleviate suffering.

Throughout the study, several measures were implemented to alleviate animal suffering, including daily health monitoring, maintenance of appropriate housing conditions, provision of adequate nutrition and hydration, and minimization of unnecessary handling. Animals exhibiting severe distress, persistent respiratory difficulty, excessive weight loss, or signs of severe pain were designated for immediate humane intervention or euthanasia according to predefined humane endpoint criteria.

#### 3.6.4. Toxicological Assessment Parameters

To evaluate the potential systemic and organ-specific toxicity of mosquito repellent exposure, a comprehensive set of clinical, physiological, hematological, biochemical, and histopathological parameters was assessed in the experimental animals (Fradin, 1998).

**(a)** Clinical Observations

Animals were monitored daily throughout the exposure period to assess overall health and detect any behavioral or physiological changes. Systematic observations included changes in activity level, grooming behavior, and signs of lethargy, as well as indicators of respiratory distress, including dyspnea, nasal irritation, and abnormal breathing patterns. In addition, animals were examined for evidence of skin irritation or dermal abnormalities, and their daily food and water intake was recorded to assess potential effects on metabolic function. Collectively, these observations served as early indicators of acute or sub-chronic toxic responses associated with exposure to mosquito repellent formulations.

**(b)** Body Weight and Organ Index Analysis

The body weight of all experimental animals was measured weekly using a digital weighing scale to monitor growth patterns and systemic health. At the termination of the study, animals were euthanized under ethical guidelines, and major vital organs, including the lungs, liver, and kidneys, were carefully excised, blotted, and weighed. Organ-to-body weight ratios (organ indices) were calculated to assess potential organ enlargement or atrophy as indicators of toxicity. Alterations in these indices were considered reflective of physiological stress or organ-specific damage induced by repellent exposure.

Organ-to-body weight ratio (organ index) is a biometric metric calculated as

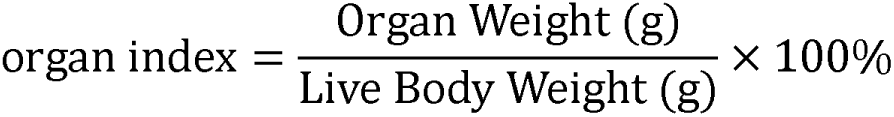

It normalizes organ weights across varying body sizes, making it a highly sensitive biomarker used in toxicology, veterinary medicine, and biological research to detect early-stage organ enlargement, atrophy, or toxicity

**(c)** Hematological Analysis

Blood samples were collected using standard laboratory procedures for hematological evaluation. Key parameters analyzed included hemoglobin concentration (Hb), total leukocyte count (TLC) using hematology analyzers (FUNKEEP MX-60). These measurements were used to assess the systemic immune response, determine inflammatory status, and identify potential hematopoietic disturbances resulting from exposure to mosquito repellent formulations.

**(d)** Biochemical Parameters

Serum biochemical analysis was performed to assess hepatic and renal functional integrity using standard biochemistry analyser (Sinothinker SK3002B1). Liver function was evaluated by measuring alanine aminotransferase (ALT) and aspartate aminotransferase (AST), while kidney function was assessed through serum creatinine and blood urea levels. Variations in these parameters were interpreted as indicators of potential hepatotoxicity and nephrotoxicity associated with chronic exposure to mosquito repellent agents.

**(e)** Histopathological Examination

At the end of the exposure period, tissue samples from the lungs, liver, and skin were collected and fixed in 10% neutral buffered formalin. The samples underwent standard histological processing, followed by sectioning and staining using Hematoxylin and Eosin (H&E). Microscopic evaluation was conducted to identify inflammatory cell infiltration, cellular degeneration or necrosis, and structural or morphological alterations in tissue architecture. The histopathological findings provided definitive evidence of organ-level toxicity and complemented the biochemical and clinical observations, enabling a comprehensive assessment of the effects of mosquito repellent exposure (Fradin, 1998).

### 3.7. Statistical analysis

Experimental data from laboratory tests, field evaluations, safety assessments, and consumer surveys were analyzed using appropriate statistical software. Descriptive statistics were used to summarize the data and present the overall distribution of measured variables. Comparative analyses among different mosquito repellent products and study groups were performed using one-way analysis of variance (ANOVA) and independent-samples t-tests, as appropriate. All results were expressed as mean ± standard deviation (SD). Statistical significance was determined at a confidence level of p < 0.05, which was considered indicative of a statistically significant difference between the study variables.

## 4. Result and Discussion

### 4.1. The Choice of Mosquito Species

To evaluate the repellent activity against mosquitoes, we conducted preliminary experiments with the common mosquito species Culex pipiens, Aedes togoi, and Aedes albopictus. Culex pipiens, the common house mosquito, however, is not ideal for the repellency test in a laboratory setting because it feeds on humans only at night due to its nocturnal habits. On the other hand, Aedes togoi showed much less biting activity than Aedes albopictus throughout the experimental setting, which is not optimal for quantifying the biting rate to assess the effect of repellents. Thus, Aedes albopictus was chosen to evaluate the effect of repellent activities clearly in the experimental setting.

### 4.2. Patch Test for Mosquito Repellents

DEET was tested on 6 volunteers. Initial skin tests on volunteers’ forearms were performed to assess their allergic responses to the three repellents. As determined by a dermatologist, none of the volunteers had allergic reactions after 8h (data not shown).

### 4.3. Repellent Effect for DEET, Coils, Vaporizers, and Aerosol, Natural Repellent

As hazards from mosquitoes have gradually increased, many kinds of mosquito repellents have been manufactured to protect humans against mosquito bites. Because mosquito repellents have played an important role in protecting humans from vector-borne diseases caused by mosquitoes, standardized guidelines are needed to evaluate their efficacy. In the United States, for example, repellents are tested against mosquitoes and other pests according to the guidelines of the Environmental Protection Agency (US EPA (Yoon et al., 2015)) and the American Society for Testing and Materials (ASTM). Although European guidelines have not been developed, the efficacy of these repellents has been tested according to the World Health Organization Pesticide Evaluation Scheme (WHOPES) and the US EPA guidelines, which are considered the international standard testing guidelines.

The repellent and insecticidal efficacy of the evaluated products, comprising a topical DEET-based cream, various commercial mosquito coils, liquid vaporizers, aerosols, and a natural plant-based repellent spray, was systematically evaluated through immediate knockdown action, 24-hour post-exposure mortality, and human-landing/biting protection assays.

#### 4.3.1. Immediate Knockdown Efficacy (10-Minute Exposure)

The immediate knockdown performance of 14 household repellent formulations was assessed 10 minutes post-exposure using a sample size of 100 mosquitoes per trial across three independent replicates. The mean knockdown rates varied significantly by product brand and formulation (table 2).

**Table 2:**
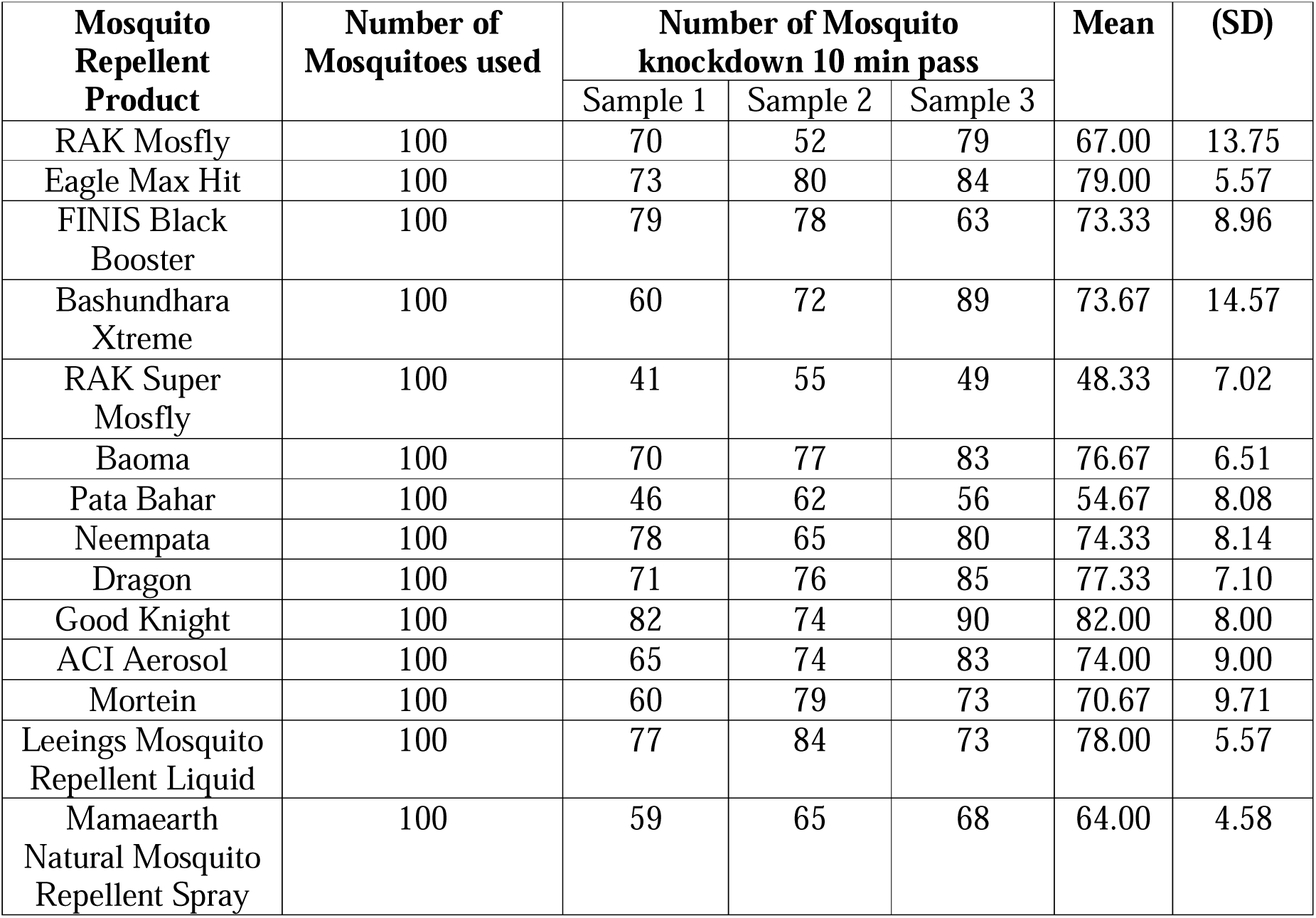
Number of Mosquito knockdown after 10 min of product use.

High-Performing Formulations: Good Knight (vaporizer/coil formulation) demonstrated the highest immediate knockdown efficacy with a mean of 82.00 ±8.00 mosquitoes, closely followed by Eagle Max Hit 79.00 ±5.57 and Leeings Mosquito Repellent Liquid 78.00 ±5.57. Moderate-Performing Formulations: Moderate knockdown action was recorded for Dragon 77.33 ±7.10, Baoma 76.67 ±6.51, Neempata 74.33 ±8.14, ACI Aerosol 74.00 ±9.00, Bashundhara Xtreme 73.67 ±14.57, and FINIS Black Booster 73.33 ±8.96.

Low-Performing Formulations: The lowest immediate knockdowns were observed in RAK Super Mosfly 48.33 ±7.02 and Pata Bahar 54.67 ±8.08. The plant-based formulation, Mamaearth Natural Mosquito Repellent Spray, showed a modest immediate knockdown rate of 64.00 ±4.58.

To determine if these differences in immediate knockdown efficacy were statistically significant across the 14 products, a One-Way Analysis of Variance (ANOVA) was conducted (table 3). The analysis revealed a highly significant difference in knockdown rates between the product groups: F(13, 28) = 3.55, p = 0.0025

**Table 3:**
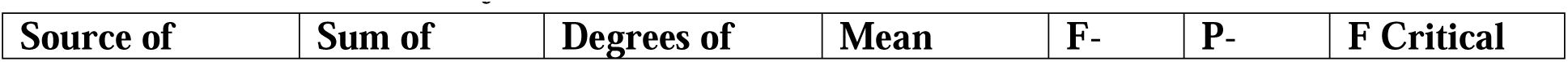
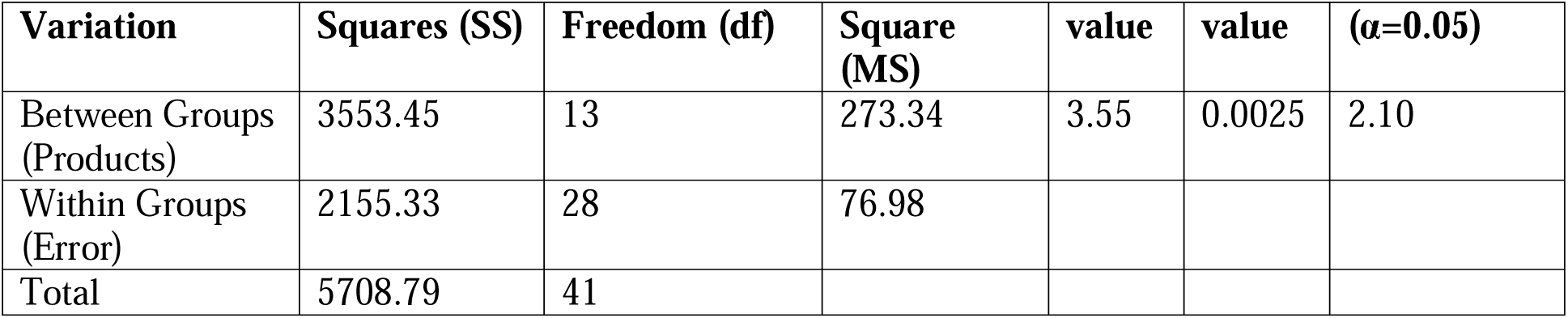
ANOVA Summary Table (Knockdown)

| Source of | Sum of | Degrees of | Mean | F- | P- | F Critical |
| --- | --- | --- | --- | --- | --- | --- |

| Variation | Squares (SS) | Freedom (df) | Square (MS) | value | value | ($\alpha=0.05$) |
| --- | --- | --- | --- | --- | --- | --- |
| Between Groups (Products) | 3553.45 | 13 | 273.34 | 3.55 | 0.0025 | 2.10 |
| Within Groups (Error) | 2155.33 | 28 | 76.98 |  |  |  |
| Total | 5708.79 | 41 |  |  |  |  |

Because the calculated F-value (3.55) significantly exceeded the critical F-value (2.10) at α = 0.05, the null hypothesis was rejected, confirming that immediate knockdown efficacy is highly dependent on the choice of repellent product.

One-Way ANOVA for 10-Minute Mosquito Knockdown

This test analyzes the differences in immediate knockdown performance among the 14 products across their three samples.

#### 4.3.2. Secondary Lethal Efficacy (24-Hour Post-Exposure Mortality)

To evaluate the long-term insecticidal potential of the vaporized, sprayed, and burned products, the mortality rate of the exposed mosquitoes was recorded 24 hours post-exposure (table 4).

**Table 4:**
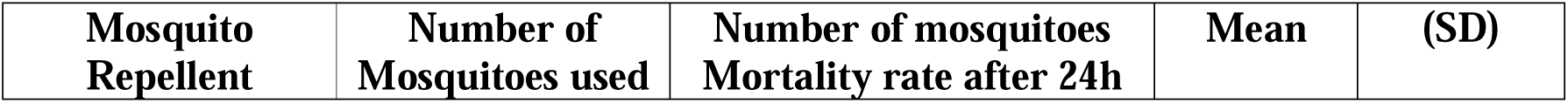
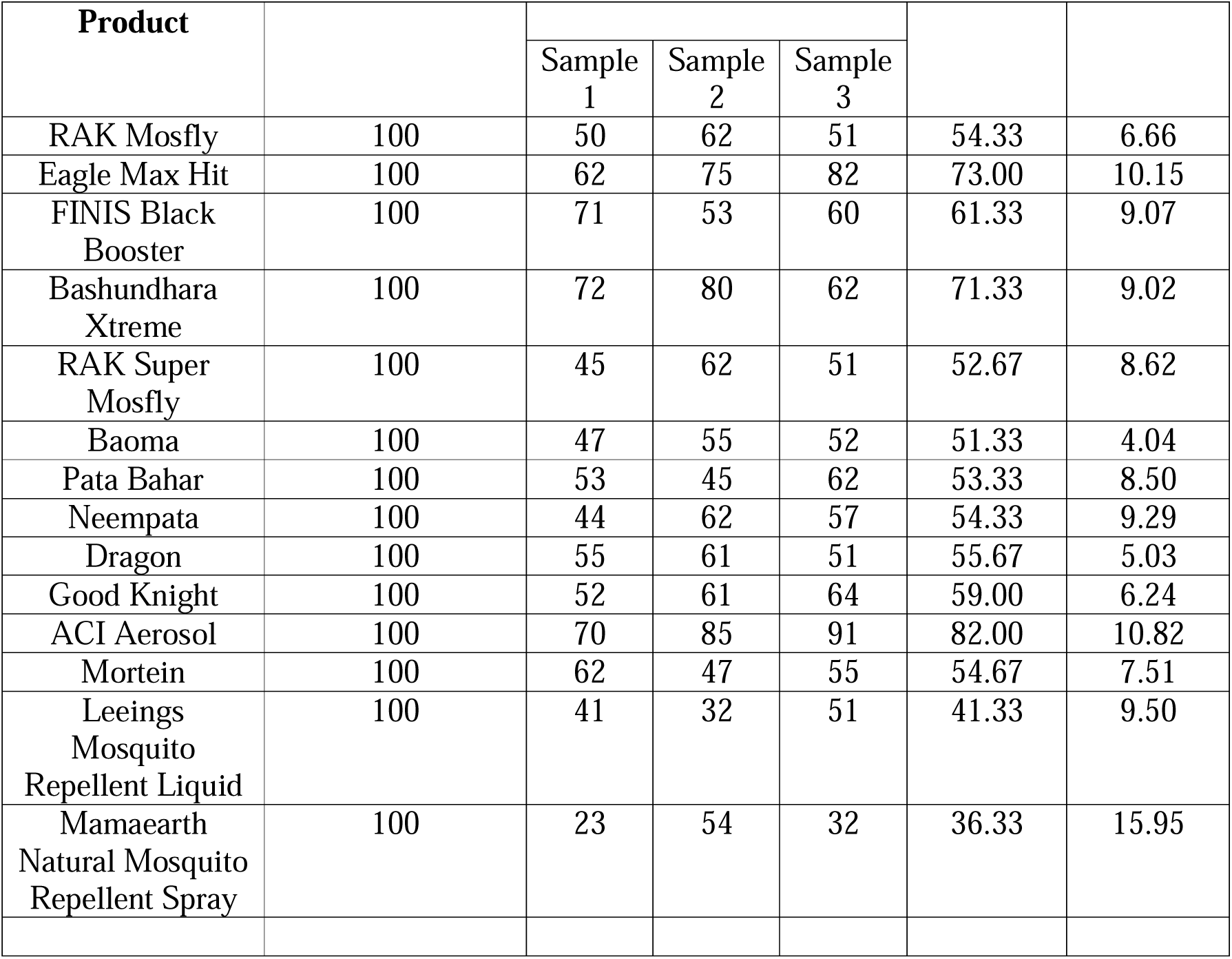
mosquito Mortality rate recorded after 24 hours.

### High Lethal Efficacy

ACI Aerosol (representing the aerosol category) yielded the highest 24-hour mortality rate, achieving a mean of 82.00 ±10.82 deaths per 100 mosquitoes. This was followed by Eagle Max Hit (73.00 ±10.15) and Bashundhara Xtreme (71.33 ±9.02).

### Moderate Lethal Efficacy

Steady insecticidal action was observed for FINIS Black Booster (61.33 ±9.07), Good Knight (59.00 ±6.24), Dragon (55.67 ±5.03), Mortein (54.67 ±7.51), RAK Mosfly (54.33 ±6.66), and Neempata (54.33 ±9.29).

### Low Lethal Efficacy

In contrast, the liquid vaporizer Leeings Mosquito Repellent Liquid (41.33 ±9.50) and the plant-based Mamaearth Natural Mosquito Repellent Spray (36.33 ±15.95) exhibited the lowest secondary lethal efficacy, indicating that their primary mode of action is deterrent or repellency-based rather than insecticidal.

The statistical significance of these mortality variations was validated using a second One-Way ANOVA (table 5): F(13, 28) = 5.28, p = 0.0001

**Table 5:**
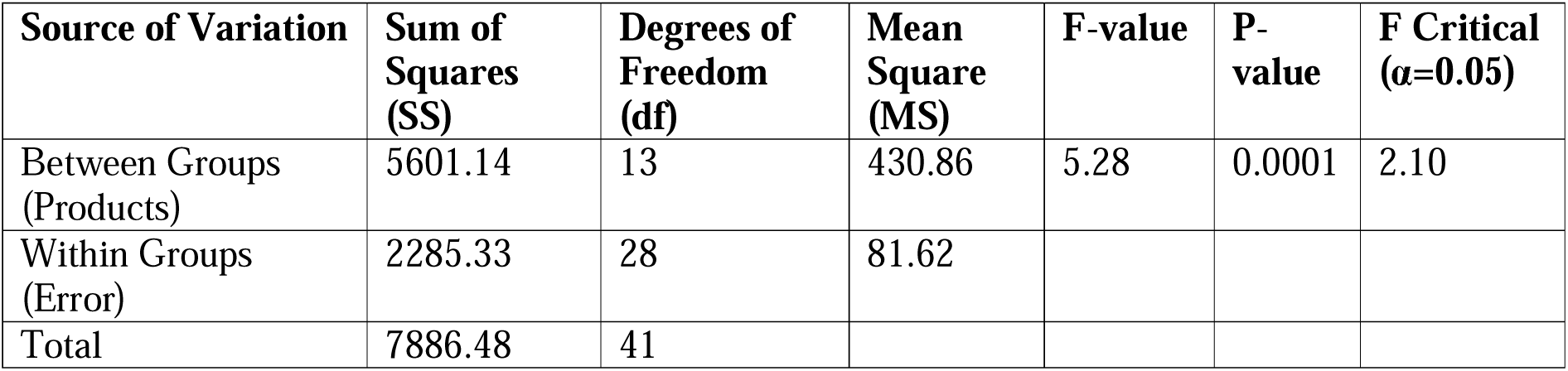
ANOVA Summary Table (Mortality Rate)

The calculated F-value (5.28) is substantially greater than the critical value (2.10) with an extremely small p-value (p < 0.001), demonstrating that the secondary lethal efficacy and overall mortality rates differ highly significantly among the tested repellent classes.

One-Way ANOVA for 24-Hour Mosquito Mortality Rate

This test analyzes the differences in long-term lethal efficacy among the 14 products across their three samples.

#### 4.3.3. Personal Repellent Efficacy of DEET-Based Positive Control

For personal topical protection, a standard cream formulation containing DEET (Dabur Odomos Cream) was evaluated on 6 human volunteers to establish baseline landing and bite prevention metrics under controlled exposure (table 6).

**Table 6:**
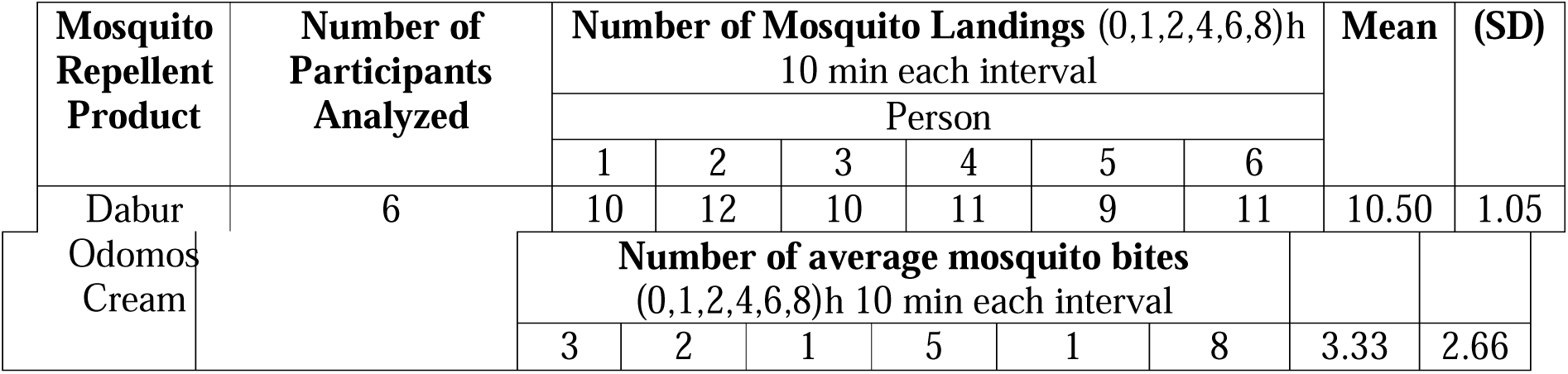
number of mosquito Landings (0,1,2,4,6,8)h 10 min each interval and Number of average mosquito bites (0,1,2,4,6,8)h 10 min each interval.

Over the cumulative evaluation intervals spanning 0, 1, 2, 4, 6, and 8 hours, the DEET-based positive control successfully limited the mean number of mosquito landings to 10.50 ±1.05 landings per 10-minute interval per volunteer. Under the same multi-hour observation window, the mean number of confirmed mosquito bites was successfully suppressed to 3.33 ±2.66 per volunteer. These results demonstrate that while household vaporizers, aerosols, and coils are highly effective at spatial knockdown and environmental mortality, topical DEET applications provide an essential personal barrier that successfully minimizes physical host-vector contact.

### 4.4. Safety and Toxicological Evaluation of Mosquito Repellent Products in a Rat Model

#### 4.4.1 Clinical Observations

Rats exposed to mosquito coil smoke exhibited mild respiratory distress, reduced activity, and occasional nasal irritation. Vaporizer and aerosol-exposed animals showed minimal behavioral changes. No significant abnormalities were observed in the DEET and natural repellent groups.

#### 4.4.2 Body Weight and Organ Index

Weekly body weight gain of rats across different exposure groups. Coil, vaporizer and aerosol exposed animals showed a significant reduction in weight gain compared to controls (p < 0.05), indicating systemic stress (Figure 1).

**Figure 1:**
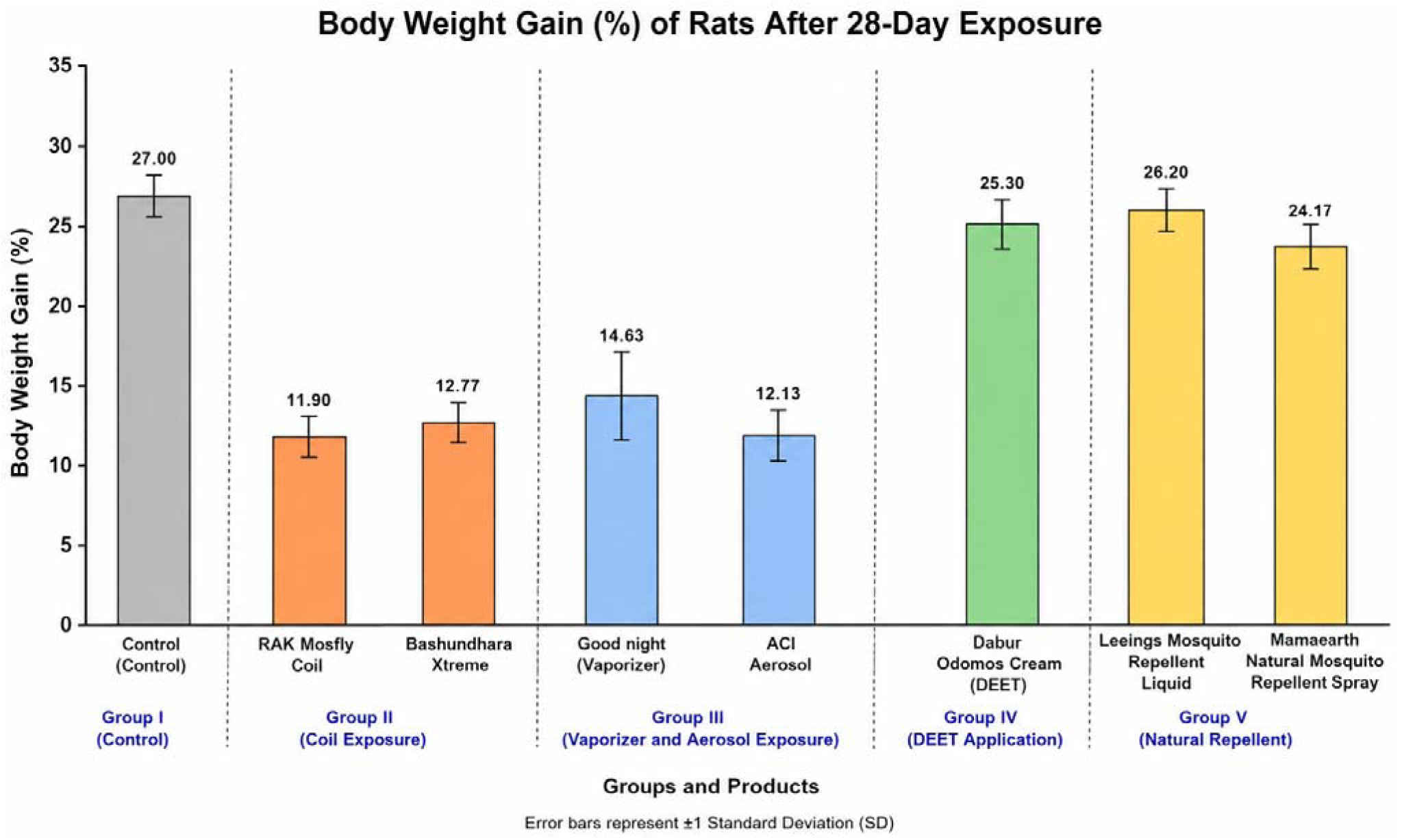
Body Weight Gain across Experimental Groups

The comparative analysis of the lung organ index demonstrated distinct differences among the experimental groups, reflecting the pulmonary effects of different mosquito repellent products (Figure 2). The control group exhibited a normal lung organ index of 0.59 ± 0.01%. In contrast, the highest lung organ indices were observed in the mosquito coil exposure groups, with RAK Mosfly Coil (0.85 ± 0.02%) and Bashundhara Xtreme (0.84 ± 0.03%), indicating marked pulmonary enlargement consistent with respiratory inflammation and tissue injury following prolonged inhalation exposure. The vaporizer and aerosol groups also showed elevated lung organ indices, with ACI Aerosol (0.76 ± 0.02%) producing a greater increase than Good Night (0.71 ± 0.02%), suggesting moderate pulmonary stress compared with the coil-exposed groups. In comparison, the DEET-based repellent (Dabur Odomos cream) produced only a slight increase in lung organ index (0.63 ± 0.02%), remaining close to the control value. Similarly, the natural repellent groups, Leeings mosquito repellent liquid (0.59 ± 0.01%) and Mamaearth Natural Mosquito Repellent Spray (0.59 ± 0.02%), exhibited lung organ indices comparable to the control group, indicating no detectable adverse effects on lung morphology under the experimental conditions. Overall, these findings suggest that mosquito coil smoke induced the greatest pulmonary toxicity, vaporizer and aerosol exposure caused moderate respiratory effects, whereas DEET-based and natural repellents produced minimal or negligible alterations in lung organ index.

**Figure 2:**
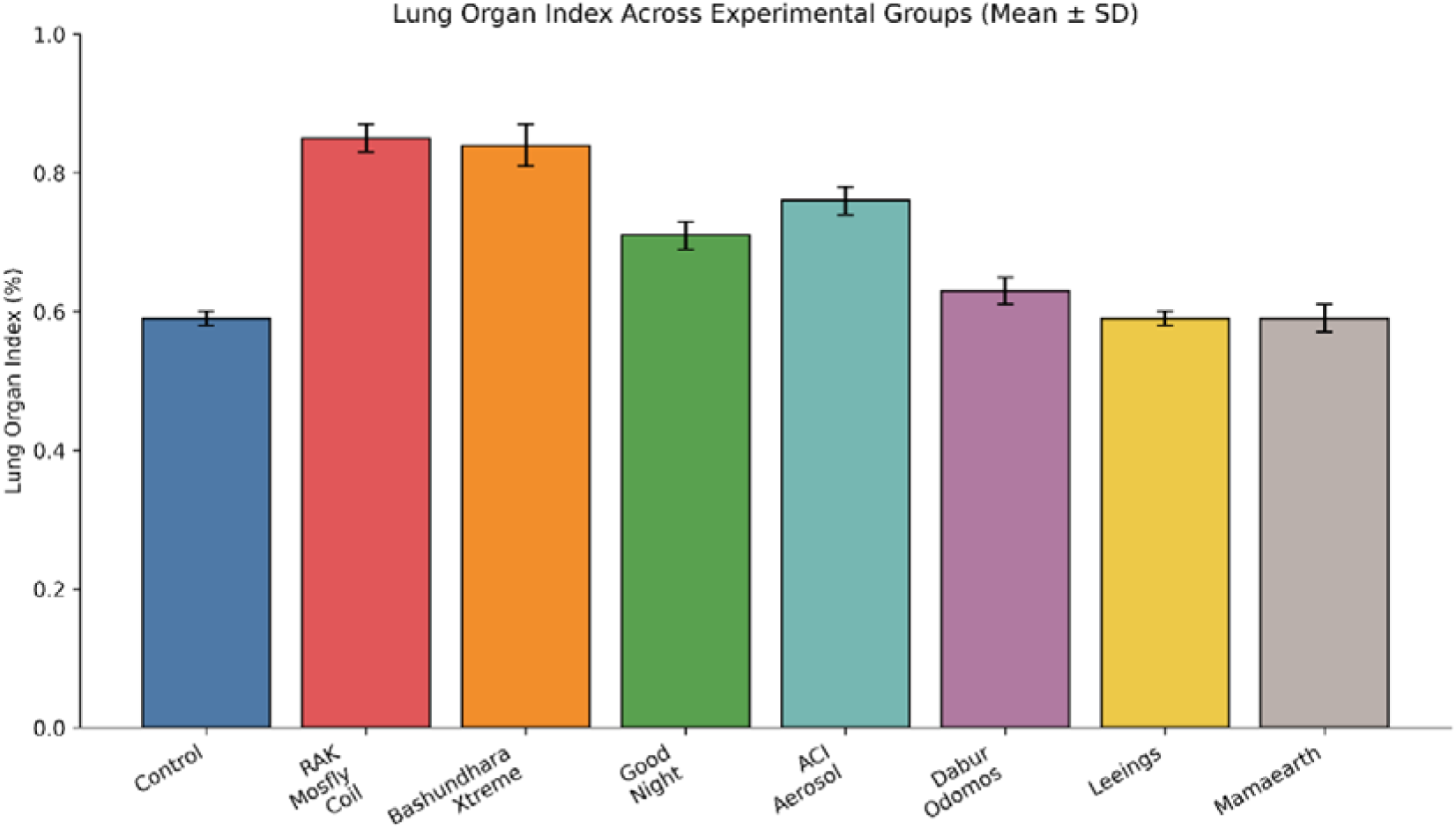
Lung Organ Index across Experimental Groups

#### 4.4.3 Hematological Findings

The total leukocyte count (TLC) varied markedly among the experimental groups, reflecting the inflammatory response induced by different mosquito repellent products (Figure 3). The control group exhibited a normal TLC of 8070 ± 122.1 cells/µL. In contrast, the mosquito coil exposure groups showed the highest leukocyte counts, with RAK Mosfly Coil recording 11850 ± 272 cells/µL and Bashundhara Xtreme recording 11550 ± 158.3 cells/µL, indicating a pronounced systemic inflammatory response following prolonged inhalation of coil smoke. The vaporizer and aerosol groups also demonstrated elevated TLC values compared with the control group. Exposure to ACI Aerosol resulted in a TLC of 10789 ± 512 cells/µL, while Good Night produced a TLC of 9752 ± 126.8 cells/µL, suggesting a moderate inflammatory response that was less severe than that observed with mosquito coil exposure. In comparison, the DEET-based repellent (Dabur Odomos) produced only a slight increase in TLC (8859 ± 325.5 cells/µL) relative to the control group. Similarly, the **natural repellent formulations, Leeings mosquito repellent (8157 ± 221.7 cells/µL) and Mamaearth Natural Mosquito Repellent Spray (8105 ± 106.6 cells/µL), exhibited leukocyte counts that were comparable to the control group, indicating minimal or no detectable inflammatory response under the experimental conditions.

**Figure 3:**
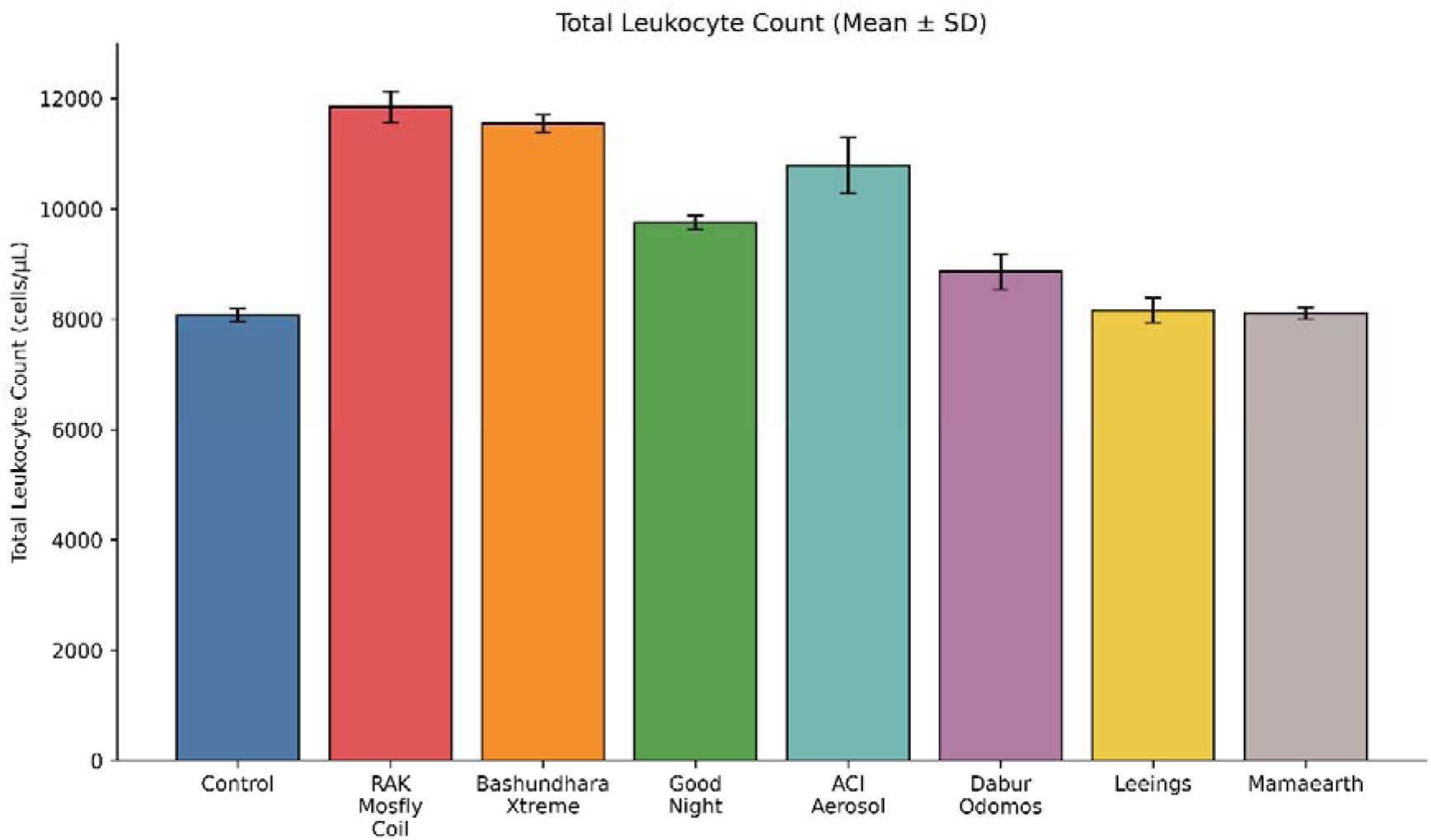
Total Leukocyte Count (TLC) Across Experimental Groups

The hemoglobin (Hb) levels varied among the experimental groups, reflecting the differential effects of mosquito repellent exposure on hematological status (Figure 4). The control group exhibited a normal hemoglobin concentration of 13.4 ± 0.30 g/dL. In contrast, the mosquito coil exposure groups demonstrated the greatest reduction in Hb levels, with RAK Mosfly Coil recording 9.80 ± 0.68 g/dL and Bashundhara Xtreme recording 9.71 ± 0.56 g/dL, suggesting that prolonged inhalation of mosquito coil smoke adversely affected erythrocyte-related parameters and oxygen-carrying capacity. The vaporizer and aerosol groups also showed decreased hemoglobin concentrations compared with the control group. Exposure to ACI Aerosol resulted in an Hb level of 9.56 ± 0.72 g/dL, representing the lowest value among all treatment groups, while Good Night exhibited a relatively moderate reduction (11.06 ± 0.25 g/dL). These findings indicate that exposure to vaporizer and aerosol emissions may also impair hematological function, although the magnitude of the effect was generally lower than that observed with mosquito coil exposure. In comparison, the **DEET-based repellent (Dabur Odomos) maintained a hemoglobin level of 13.05 ± 0.56 g/dL, which was comparable to that of the control group. Likewise, the natural repellent formulations, Leeings (13.60 ± 0.39 g/dL) and Mamaearth Natural Mosquito Repellent Spray (13.50 ± 0.26 g/dL), exhibited hemoglobin concentrations similar to or slightly higher than the control value, indicating no evidence of hematological impairment following treatment.

**Figure 4:**
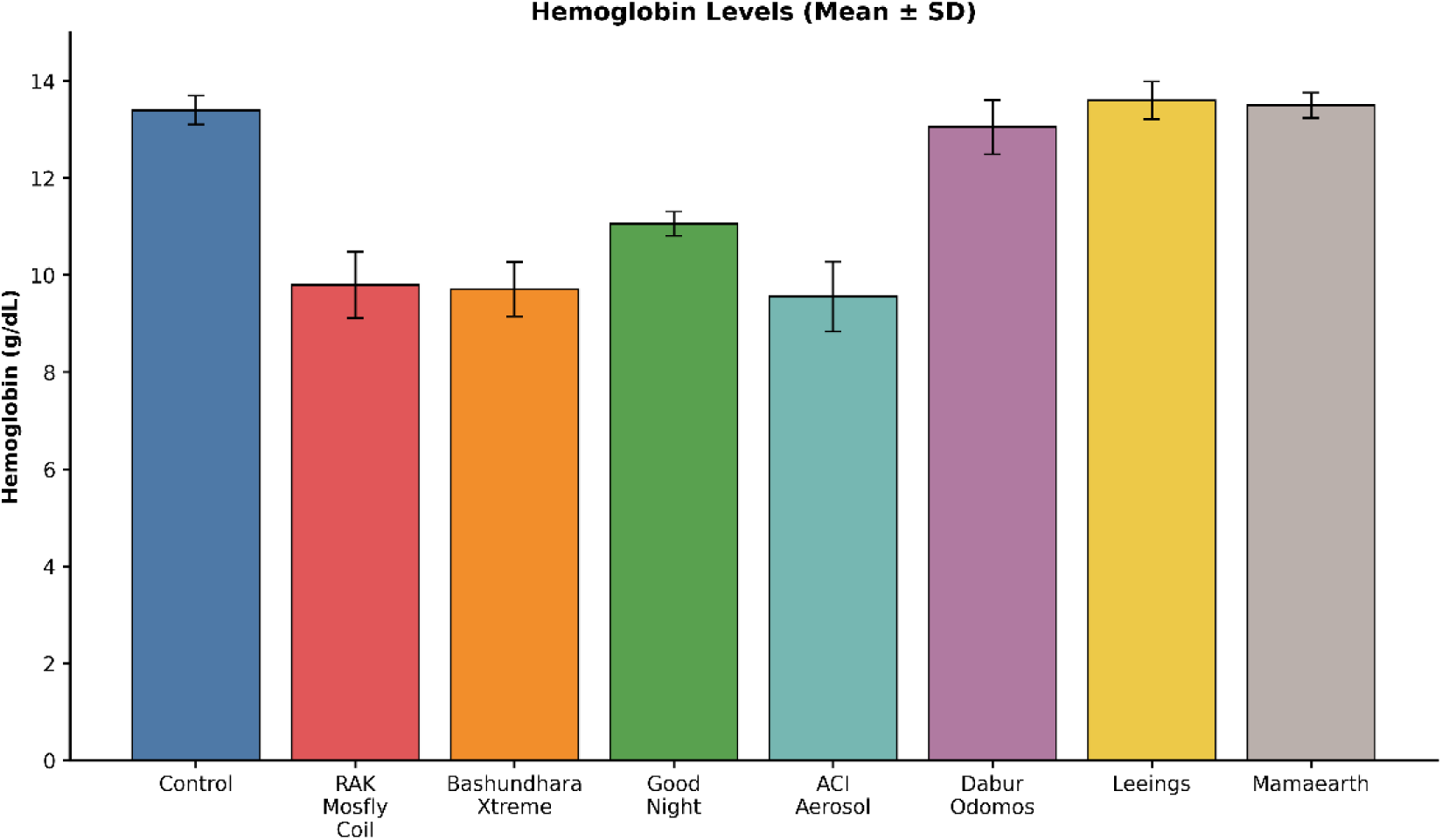
Hemoglobin Levels across Experimental Groups

#### 4.4.4 Biochemical Analysis

The biochemical analysis revealed distinct alterations in liver and kidney function biomarkers among the experimental groups following exposure to different mosquito repellent products (Figure 5). The control group exhibited normal serum levels of ALT (32 ± 1.20 U/L), AST (36 ± 1.25 U/L), creatinine (0.80 ± 0.045 mg/dL), and blood urea (28 ± 0.95 mg/dL), representing baseline physiological conditions. The mosquito coil exposure groups demonstrated the most pronounced biochemical alterations. Rats exposed to RAK Mosfly Coil showed markedly elevated ALT (68 ± 0.86 U/L) and AST (74 ± 0.86 U/L) levels, while those exposed to Bashundhara Xtreme exhibited similarly increased ALT (66 ± 1.20 U/L) and AST (75 ± 0.95 U/L) levels, indicating substantial hepatocellular injury and hepatic stress following prolonged inhalation of mosquito coil smoke. In addition, renal biomarkers were elevated in these groups, with creatinine increasing to 1.30 ± 0.05 mg/dL and 1.20 ± 0.062 mg/dL, and blood urea rising to 44 ± 1.25 mg/dL and 45 ± 0.75 mg/dL, respectively, suggesting mild impairment of renal function. The vaporizer and aerosol groups exhibited moderate biochemical changes compared with the coil groups. Exposure to Good Night resulted in ALT and AST levels of 45 ± 1.36 U/L and 48 ± 1.32 U/L, respectively, whereas ACI Aerosol produced slightly higher enzyme activities (ALT: 46 ± 1.78 U/L; AST: 51 ± 0.56 U/L). Correspondingly, serum creatinine increased to 1.00 ± 0.04 mg/dL and 1.10 ± 0.035 mg/dL, while blood urea reached 34 ± 0.90 mg/dL and 33 ± 1.80 mg/dL, indicating moderate hepatic stress accompanied by only mild renal involvement. In contrast, the DEET-based repellent (Dabur Odomos cream) produced only minimal biochemical alterations, with ALT (34 ± 1.35 U/L), AST (37 ± 1.21 U/L), creatinine (0.82 ± 0.071 mg/dL), and blood urea (29 ± 2.30 mg/dL) remaining close to the control values. Likewise, the natural repellent formulations, Leeings mosquito repellent liquid (ALT: 31 ± 0.96 U/L; AST: 35 ± 0.74 U/L; creatinine: 0.79 ± 0.05 mg/dL; blood urea: 27 ± 0.72 mg/dL) and Mamaearth Natural Mosquito Repellent Spray (ALT: 32 ± 0.82 U/L; AST: 36 ± 0.56 U/L; creatinine: 0.78 ± 0.04 mg/dL; blood urea: 27 ± 1.10 mg/dL), exhibited biochemical profiles comparable to those of the control group, indicating negligible hepatic or renal toxicity under the experimental conditions.

**Figure 5:**
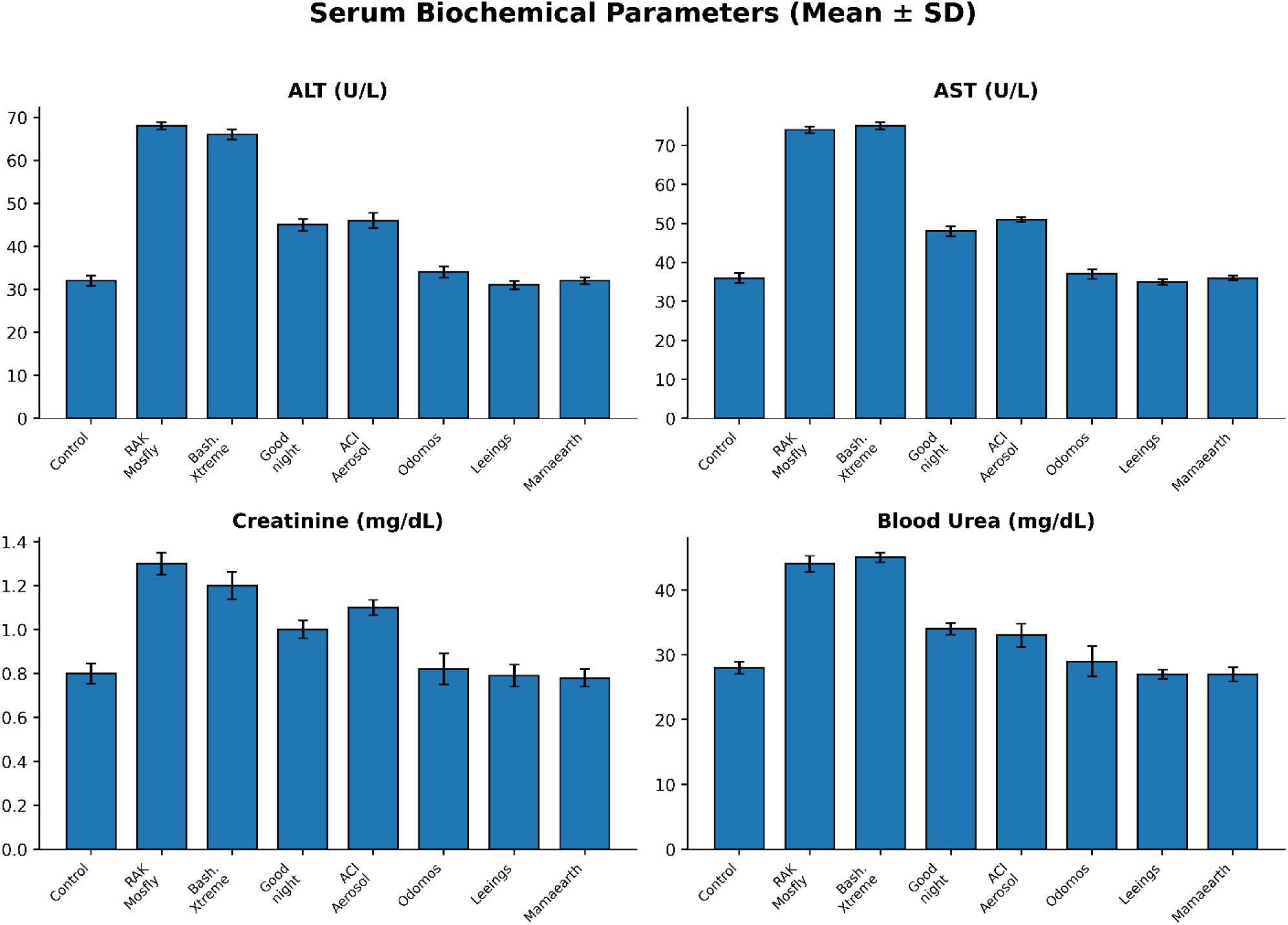
The biochemical analysis of ALT, AST, Creatinine, and Blood Urea in control, coil, vaporizer, aerosol, DEET, and natural group

#### 4.4.5 Histopathology

Histopathological examination of H&E-stained lung, liver, and skin tissues revealed marked differences in tissue responses among the experimental groups following exposure to various mosquito repellent products (Figures 6 and 7). The control group exhibited normal tissue architecture in all examined organs, characterized by intact alveolar structures in the lungs, well-organized hepatic cords with normal central veins in the liver, and a normal epidermal and dermal architecture in the skin, with no evidence of inflammatory cell infiltration or structural abnormalities. Correspondingly, all histopathological severity scores were 0 for inflammation, epithelial damage, congestion, and tissue alteration.

**Figure 6:**
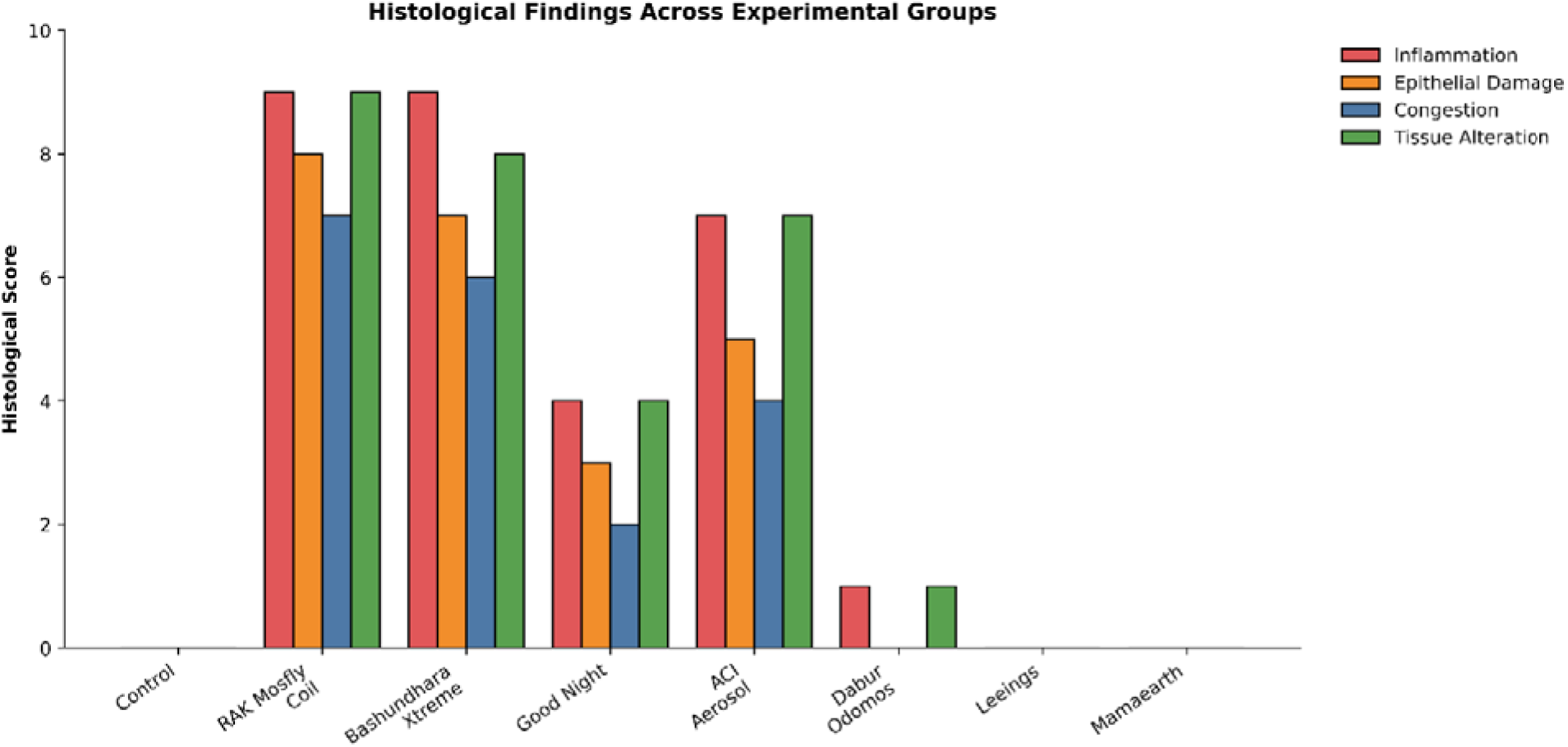
The histopathological analysis graph has been created based on the inflammation, epithelial cell damage, and congestion, tissue alteration on control, coil, vaporizer, aerosol, DEET, and natural product exposure.

**Figure 7:**
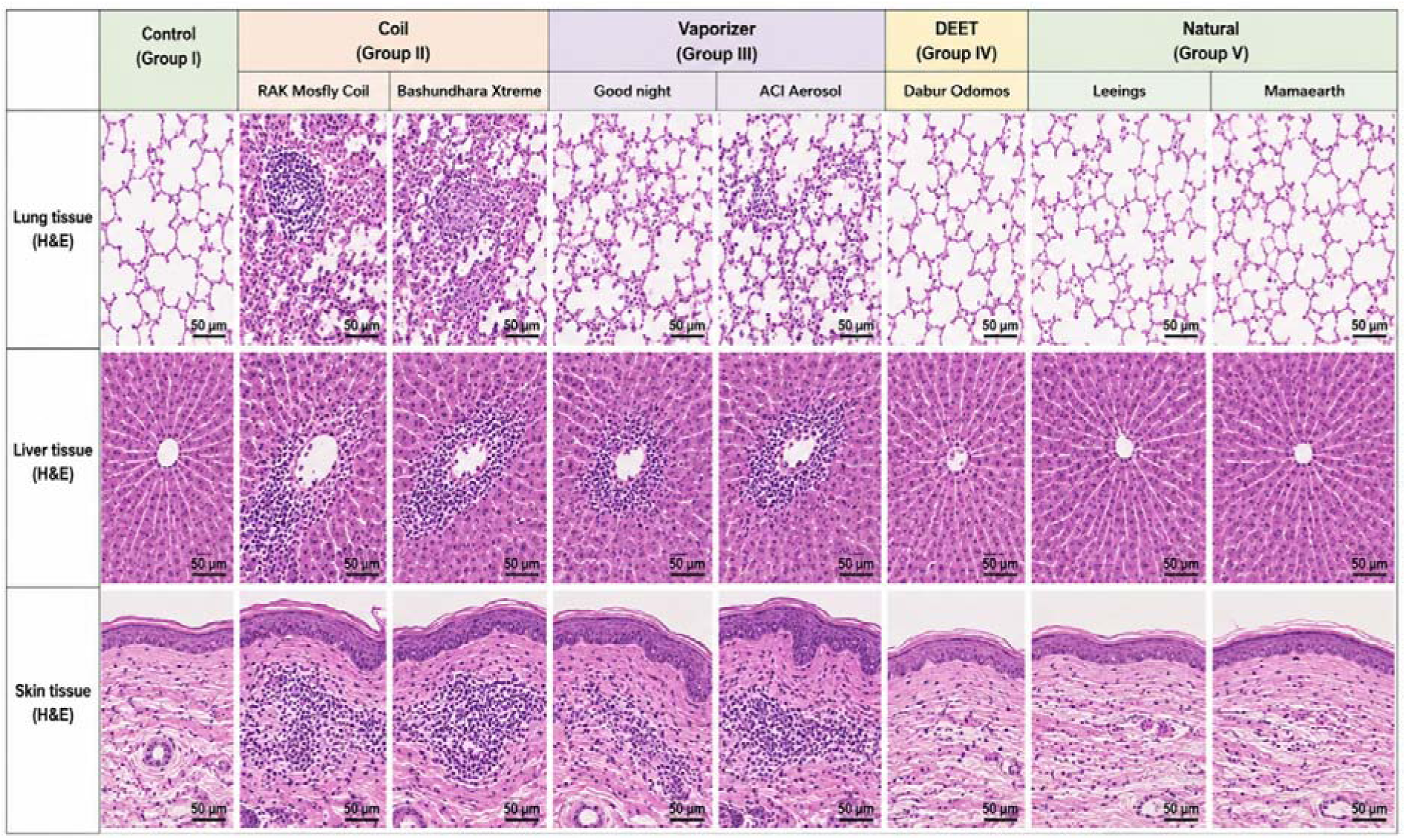
H&E-stained section (40X). Control and natural group (Leeings, mamaearth) show normal histoarchitecture with minimal inflammation. Coil (RAK Mosfly coil, Bashundhara Xtreme) shows severe inflammatory cell infiltration, epithelial damage, congestion, and tissue alteration. Vaporizer (Good Night, ACI Aerosol) shows moderate changes, DEET group (Dabur Odomos) shows mild changes.

The mosquito coil exposure groups demonstrated the most severe histopathological changes. Rats exposed to RAK Mosfly Coil exhibited the highest pathological scores, including inflammation (9/10), epithelial damage (8/10), congestion (7/10), and tissue alteration (9/10). Similarly, Bashundhara Xtreme produced severe lesions with inflammation (9/10), epithelial damage (7/10), congestion (6/10), and tissue alteration (8/10). Microscopic examination revealed extensive inflammatory cell infiltration, disruption of alveolar architecture, epithelial degeneration, vascular congestion, and marked structural alterations in lung tissue. In the liver, these groups showed prominent periportal inflammatory infiltration, hepatocellular disorganization, and vascular congestion, while the skin exhibited epidermal thickening, inflammatory cell infiltration, and dermal structural disruption. These findings indicate substantial tissue injury following prolonged exposure to mosquito coil smoke.

The vaporizer and aerosol groups displayed moderate histopathological alterations. Exposure to Good Night resulted in inflammation (4/10), epithelial damage (3/10), congestion (2/10), and tissue alteration (4/10), whereas ACI Aerosol produced comparatively greater tissue injury with scores of 7/10, 5/10, 4/10, and 7/10, respectively. Histological examination demonstrated mild-to-moderate inflammatory infiltration, partial epithelial disruption, and localized vascular congestion, while the overall tissue architecture remained largely preserved compared with the coil-exposed groups.

In contrast, the DEET-based repellent (Dabur Odomos) exhibited only minimal histopathological changes, with inflammation (1/10), epithelial damage (0/10), congestion (0/10), and tissue alteration (1/10). H&E sections showed largely normal lung, liver, and skin morphology with only occasional inflammatory cells and no evidence of significant tissue degeneration, suggesting minimal toxicity under the conditions of this study.

The natural repellent formulations, Leeings mosquito repellent and Mamaearth Natural Mosquito Repellent Spray, demonstrated no detectable histopathological abnormalities, with severity scores of 0 for all evaluated parameters. Microscopic examination confirmed normal pulmonary alveoli, intact hepatic lobular architecture, and healthy skin morphology without inflammatory infiltration, epithelial injury, vascular congestion, or structural tissue alteration. Their histological appearance was indistinguishable from that of the control group.

Overall, the histopathological findings demonstrate a clear toxicity gradient among the tested mosquito repellent products. Mosquito coil smoke produced the most severe pathological alterations, characterized by extensive inflammation, epithelial injury, vascular congestion, and structural tissue damage across multiple organs. Vaporizer and aerosol exposure induced moderate lesions, with ACI Aerosol producing greater tissue damage than Good Night. In contrast, the DEET-based repellent caused only minimal histological changes, while the natural repellents (Leeings and Mamaearth Natural Mosquito Repellent Spray) maintained normal tissue architecture comparable to the control group. These observations are consistent with the biochemical and hematological findings, indicating that combustion-based mosquito repellents pose the greatest toxicological risk, whereas topical DEET-based and natural repellent formulations exhibit substantially lower tissue toxicity under the experimental conditions. (Diaz, 2016).

## 5. Recommendations for users

Based on the findings of this study, consumers should carefully select mosquito repellent products by considering both their effectiveness against mosquitoes and their potential health effects (Peterson et al., 2006). Prolonged exposure to mosquito coil smoke should be minimized, particularly in poorly ventilated indoor environments, as the present study demonstrated that coil exposure produced the greatest toxicological effects, including reduced body weight gain, increased lung organ index, elevated total leukocyte count, decreased hemoglobin levels, altered liver and kidney biochemical markers, and severe histopathological damage in the lung, liver, and skin tissues (J et al., 2025; Peng et al., 2022a). When indoor protection is required, liquid vaporizers and aerosols may be preferred over mosquito coils because they produced comparatively lower toxicological effects, although adequate room ventilation should always be maintained to reduce the accumulation of airborne chemicals and minimize respiratory exposure (Norris & Coats, 2017). For outdoor activities or situations requiring prolonged personal protection, DEET-based topical repellents are recommended, as they maintained hematological, biochemical, and histopathological parameters close to those of the control group, indicating a favorable safety profile when used according to the manufacturer’s instructions. Similarly, natural repellents such as **Leeings Mosquito Repellent** and **Mamaearth Natural Mosquito Repellent Spray** demonstrated negligible systemic toxicity and preserved normal organ structure and function, making them suitable alternatives for routine household use, particularly for children, pregnant women, elderly individuals, and people with respiratory sensitivity (Peterson et al., 2006). Regardless of the product used, consumers should strictly follow the recommended dosage and application instructions and avoid excessive or prolonged exposure (Hazarika & Krishnatreyya, 2025). Furthermore, mosquito repellents should be used as part of an integrated mosquito control strategy that includes eliminating mosquito breeding sites, using insecticide-treated bed nets, installing window screens, and maintaining proper environmental sanitation. Adopting these practices will help maximize protection against mosquito-borne diseases while minimizing unnecessary chemical exposure and associated health risks.

## 6. Conclusion

This study provides a comprehensive comparative evaluation of the efficacy and toxicological safety of commercially available mosquito repellent products used in Bangladesh. The results demonstrated that mosquito coils, although effective and affordable, posed the greatest health risks, causing significant physiological, hematological, biochemical, and histopathological alterations following prolonged exposure. Liquid vaporizers and aerosols exhibited moderate toxicity with comparatively lower adverse effects than mosquito coils. In contrast, DEET-based topical repellents provided effective mosquito protection while maintaining normal physiological and biochemical parameters, indicating a favorable safety profile when used according to the manufacturer’s recommendations. Similarly, the natural repellents (Leeings Mosquito Repellent and Mamaearth Natural Mosquito Repellent Spray) showed the lowest toxicological impact, with minimal changes in organ function and tissue morphology, although their protective efficacy and duration were comparatively lower (Shyam et al., 2026)..

Overall, the findings highlight a clear efficacy–safety trade-off, where combustion-based repellents provide prolonged protection at the expense of increased health risks, whereas topical and natural formulations offer safer alternatives with reduced toxicity. These results emphasize the importance of selecting mosquito repellents based not only on their effectiveness but also on their potential health effects and intended application. Furthermore, the study underscores the need for stronger regulatory oversight, improved public awareness regarding the safe use of mosquito repellents, and continued research into low-cost, environmentally friendly, and sustainable repellent technologies that can provide effective mosquito control while minimizing risks to human health. Such advancements will contribute to safer vector control strategies and support ongoing efforts to reduce the burden of mosquito-borne diseases in endemic regions.

## Data Availability

All data used here are the author's original data and open to all for used

https://clinicaltrials.gov/study/NCT07686159

## Acknowledgements

All authors would like to express their deepest gratitude to all the individuals and organizations who contributed to the completion of this research. Their support, guidance, and expertise were invaluable in evaluating of Mosquito Repellent Products: Efficacy, Safety, and Public Health Implications.

## Financial Disclosure Statement

This work was not supported by any organization.

## Competing interests

The authors declare that they have no known competing financial interests or personal relationships that could have influenced the work reported in this paper.

## Notes

### Competing Interest Statement

The authors have declared no competing interest.

### Clinical Trial

NCT07686159

### Clinical Protocols

https://pmc.ncbi.nlm.nih.gov/articles/PMC5887648/

### Author Declarations

The experiment was approved by the Institutional Review Board of the Biomedical Research and Innovation Center, Military Institute of Science and Technology, Mirpur-1216, Dhaka, Bangladesh (approval code: 2025H009, Date of Approval: 25/ 09/2025, NCT07686159).

### Summary of Updates

Manuscript's methodology and results portion improved significantly. New tests and data are included.

