## Supplementary material for "Comprehensive Evaluation of Mosquito Repellent Products: Efficacy, Safety, and Public Health Implications": Supplymentry materials

**Table 1: Comparative Analysis of Effectiveness based on the literature**

| **Product Type** | **Example Brands** | **Active ingredients** | **Protection Duration** | **Effectiveness Level** | **Environment** | **Reference** |
| --- | --- | --- | --- | --- | --- | --- |
| Coils | RAK Mosfly, Eagle Max Hit, FINIS Black Booster, Bashundhara Xtreme, Rak Super Mosfly, Baoma, Pata Bahar, Neempata, and Dragon | [Pyrethrum](https://en.wikipedia.org/wiki/Mosquito_coil), Allethrin (d-trans-allethrin), Esbiothrin, Dimefluthrin, Meperfluthrin, Metofluthrin, Prallethrin, Permethrin, Tetramethrin | 10–12 hrs | High (outdoor + indoor) | Open & closed spaces | (Abdul Ahad et al., 2010; admin, 2022; Yildiztekin, 2016) |
| Vaporizers and Aerosol | Good Knight, ACI Aerosol, and Mortein | Transfluthrin, Prallethrin, Metofluthrin, Dimefluthrin, Allethrin / d-trans-Allethrin, Esbiothrin (s-bioallethrin), Tetramethrin, Meperfluthrin | 6–8 hrs/night | High (indoor only) | Closed rooms | (Abdul Ahad et al., 2010; admin, 2022; Yildiztekin, 2016) |
| Natural Repellent | Leeings mosquito repellent liquid, Mamaearth Natural Mosquito Repellent Spray, | Para-menthane-3,8-diol (PMD), Citronella oil, Eucalyptus oil, Lemongrass oil, Geraniol, Neem oil, Camphor, Glycerol, Clove Oil, olive oil, zinger oil, Lemongrass Oil, | 4–6 hrs | Moderate (indoor only) | Closed rooms | (Hazarika et al., 2022; Peng et al., 2022b) |
| DEET Creams | Dabur Odomos cream | DEET, [Picaridin](https://www.repel.com/tips-and-articles/understanding--active-ingredients-in-mosquito-repellents) | 4–8 hrs | High | Personal protection | (Evans, 2003; Ramaswamy et al., 2003) |

#### Table 2: Number of Mosquito knockdown after 10 min of product use

| **Mosquito Repellent Product** | **Number of Mosquitoes used** | **Number of Mosquito knockdown 10 min pass** | | | **Mean** | **(SD)** |
| --- | --- | --- | --- | --- | --- | --- |
|  |  | Sample 1 | Sample 2 | Sample 3 |  |  |
| RAK Mosfly | 100 | 70 | 52 | 79 | 67.00 | 13.75 |
| Eagle Max Hit | 100 | 73 | 80 | 84 | 79.00 | 5.57 |
| FINIS Black Booster | 100 | 79 | 78 | 63 | 73.33 | 8.96 |
| Bashundhara Xtreme | 100 | 60 | 72 | 89 | 73.67 | 14.57 |
| RAK Super Mosfly | 100 | 41 | 55 | 49 | 48.33 | 7.02 |
| Baoma | 100 | 70 | 77 | 83 | 76.67 | 6.51 |
| Pata Bahar | 100 | 46 | 62 | 56 | 54.67 | 8.08 |
| Neempata | 100 | 78 | 65 | 80 | 74.33 | 8.14 |
| Dragon | 100 | 71 | 76 | 85 | 77.33 | 7.10 |
| Good Knight | 100 | 82 | 74 | 90 | 82.00 | 8.00 |
| ACI Aerosol | 100 | 65 | 74 | 83 | 74.00 | 9.00 |
| Mortein | 100 | 60 | 79 | 73 | 70.67 | 9.71 |
| Leeings Mosquito Repellent Liquid | 100 | 77 | 84 | 73 | 78.00 | 5.57 |
| Mamaearth Natural Mosquito Repellent Spray | 100 | 59 | 65 | 68 | 64.00 | 4.58 |

### One-Way ANOVA for 10-Minute Mosquito Knockdown

This test analyzes the differences in immediate knockdown performance among the 14 products across their three samples.

#### Table 3: ANOVA Summary Table (Knockdown)

| **Source of Variation** | **Sum of Squares (SS)** | **Degrees of Freedom (df)** | **Mean Square (MS)** | **F-value** | **P-value** | **F Critical (α=0.05)** |
| --- | --- | --- | --- | --- | --- | --- |
| Between Groups (Products) | 3553.45 | 13 | 273.34 | 3.55 | 0.0025 | 2.10 |
| Within Groups (Error) | 2155.33 | 28 | 76.98 |  |  |  |
| Total | 5708.79 | 41 |  |  |  |  |

#### Table 4: mosquito Mortality rate **recorded after 24 hours**

| **Mosquito Repellent Product** | **Number of Mosquitoes used** | **Number of mosquitoes Mortality rate after 24h** | | | **Mean** | **(SD)** |
| --- | --- | --- | --- | --- | --- | --- |
|  |  | Sample 1 | Sample 2 | Sample 3 |  |  |
| RAK Mosfly | 100 | 50 | 62 | 51 | 54.33 | 6.66 |
| Eagle Max Hit | 100 | 62 | 75 | 82 | 73.00 | 10.15 |
| FINIS Black Booster | 100 | 71 | 53 | 60 | 61.33 | 9.07 |
| Bashundhara Xtreme | 100 | 72 | 80 | 62 | 71.33 | 9.02 |
| RAK Super Mosfly | 100 | 45 | 62 | 51 | 52.67 | 8.62 |
| Baoma | 100 | 47 | 55 | 52 | 51.33 | 4.04 |
| Pata Bahar | 100 | 53 | 45 | 62 | 53.33 | 8.50 |
| Neempata | 100 | 44 | 62 | 57 | 54.33 | 9.29 |
| Dragon | 100 | 55 | 61 | 51 | 55.67 | 5.03 |
| Good Knight | 100 | 52 | 61 | 64 | 59.00 | 6.24 |
| ACI Aerosol | 100 | 70 | 85 | 91 | 82.00 | 10.82 |
| Mortein | 100 | 62 | 47 | 55 | 54.67 | 7.51 |
| Leeings Mosquito Repellent Liquid | 100 | 41 | 32 | 51 | 41.33 | 9.50 |
| Mamaearth Natural Mosquito Repellent Spray | 100 | 23 | 54 | 32 | 36.33 | 15.95 |

### One-Way ANOVA for 24-Hour Mosquito Mortality Rate

This test analyzes the differences in long-term lethal efficacy among the 14 products across their three samples.

#### Table 5: ANOVA Summary Table (Mortality Rate)

| **Source of Variation** | **Sum of Squares (SS)** | **Degrees of Freedom (df)** | **Mean Square (MS)** | **F-value** | **P-value** | **F Critical (α=0.05)** |
| --- | --- | --- | --- | --- | --- | --- |
| Between Groups (Products) | 5601.14 | 13 | 430.86 | 5.28 | 0.0001 | 2.10 |
| Within Groups (Error) | 2285.33 | 28 | 81.62 |  |  |  |
| Total | 7886.48 | 41 |  |  |  |  |

**Table 6: number of mosquito Landings** (0,1,2,4,6,8)h 10 min each interval and **Number of average mosquito bites** (0,1,2,4,6,8)h 10 min each interval

| **Mosquito Repellent Product** | **Number of Participants Analyzed** | **Number of Mosquito Landings** (0,1,2,4,6,8)h 10 min each interval | | | | | | **Mean** | **(SD)** |
| --- | --- | --- | --- | --- | --- | --- | --- | --- | --- |
|  |  | Person | | | | | |  |  |
|  |  | 1 | 2 | 3 | 4 | 5 | 6 |  |  |
| Dabur Odomos Cream | 6 | 10 | 12 | 10 | 11 | 9 | 11 | 10.50 | 1.05 |
|  |  | **Number of average mosquito bites**  (0,1,2,4,6,8)h 10 min each interval | | | | | |  |  |
|  |  | 3 | 2 | 1 | 5 | 1 | 8 | 3.33 | 2.66 |
